# A new ischemic grading system to aid combat extremity vascular injury decision making

**DOI:** 10.1101/2020.08.03.20166827

**Authors:** Amila Ratnayake, Viktor A. Reva, Miklosh Bala, Achala U. Jayatilleke, Sujeewa P.B. Thalgaspitiya, David S. Kauvar, Tamara J. Worlton

## Abstract

**Introduction:** In resource limited combat settings with frequent encounters of mass casualty incidents, the decision to attempt limb salvage versus primary amputation is refined over time based on experience. This experience can be augmented by grading systems and algorithms to assist in clinical decisions. Few investigators have attempted to explicitly grade limb ischemia according to clinical criteria and study the impact of limb ischemia on clinical outcome. We suggest a new ischemia grading system based on the Rutherford ischemic classification and the V. A. Kornilov classification which we adapted to apply to the combat setting. This new tool was then retrospectively applied to combat trauma patients from the Sri Lankan Civil War.

**Method:** We retrospectively queried a prospectively maintained, single surgeon registry containing 129 extremity vascular injuries managed at a Role 3 military base hospital (MBH) from 2008 December to June 2009 during the last phase of Sri Lankan Civil war. 89 patients were analyzed for early limb salvage according to the modified Kornilov extremity ischemia index (MKEII).

**Result:** According to the MKEII, subcohort analysis of C1 (viable), C2 (threatened), and C3 (irreversible) classified injuries demonstrated a statistically significant (P < 0.001) difference in limb salvage. Further statistical evaluation demonstrated injury to popliteal region (P=0.006), severe arterial injury (P=0.018) and venous injuries (P< 0.001) had statistically significant differences in distribution between C1, C2 and C3.

**Conclusion:** By application of the MKEII, combat surgeons can rapidly and correctly select and prioritize vascular injured extremities to optimally use limited resources to achieve realistic limb salvage goals. A rigid ankle was correlated with the worst index of extremity ischemia. Further investigation into this sign as an indication for primary amputation is warranted.

## Introduction

Amputation from vascular trauma remains a considerable burden in both civilian and military populations. ^1-2^ Patients with a lower extremity vascular injury, whether from blunt or penetrating cause, are at higher risk for amputation. ^6-11^ The decision to attempt limb salvage or primarily amputate continues to be a challenging task for the combat surgeon. The Mangled Extremity Severity Score (MESS) has a limited diagnostic accuracy in predicting the need for amputation in civilian practice.^3-6^ Additionally, none of the ischemic extremity severity scoring systems have ever been conclusively validated in the military trauma setting.^7^ In a dynamic battlefront, a surgeon may have to manage multiple casualties and the decision has to be made quickly whether to attempt limb salvage and therefore commit valuable resources.

In the Sri Lankan Civil War, combat casualty care was organized from Role 1 through Role 4, with increasing sophistication of care. Role 2 was the first level capable of damage control surgery. Suspected extremity vascular injuries were prioritized for transfer to Role 3 which had vascular reconstruction capability. At the Role 3, vascular injury was assessed with physical exam and doppler. Patients with confirmed or suspected vascular injury were taken to the operating theater where a four compartment fasciotomy was performed and muscle viability was assessed. If muscles were considered non-viable, a primary amputation was performed. Viable injuries were explored and revascularized. When adequate extremity perfusion was confirmed, patients who needed advanced orthopedic and plastic surgical procedures were transferred to Role 4. All the others were managed at the Role 3 until discharge criteria were met.^8^

A core parameter relevant to limb salvage is the degree of muscle ischemia at the time of presentation and the projected worsening of ischemic load during the timeline of attempted revascularization. There is a need to develop and validate objective, injury-specific assessment tools which take into account changing priorities in resource constrained military settings. (Figure 1)

**Figure 1:**
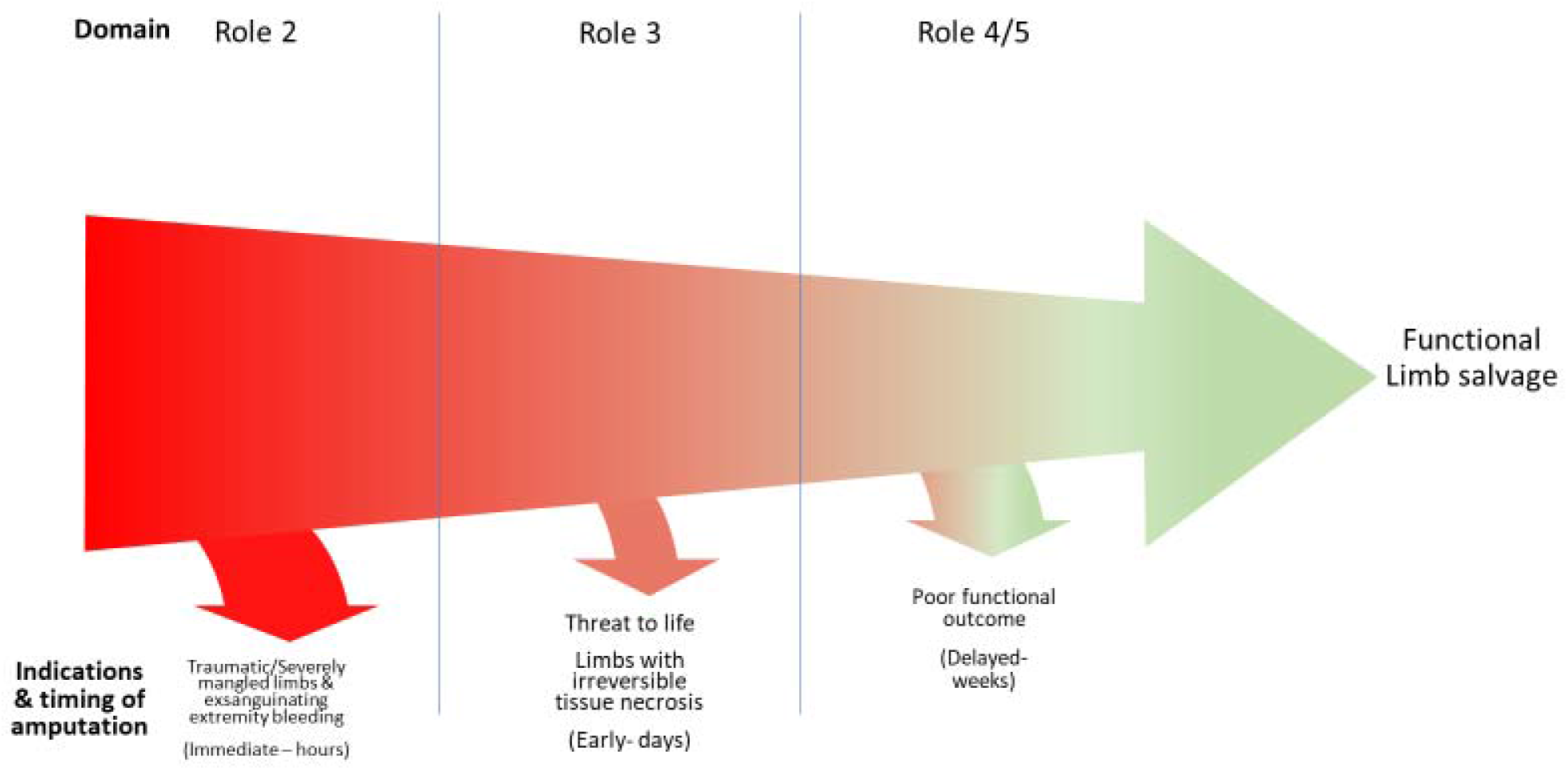
Staged amputation decisions in the combat setting

## Methods

This study had a waiver of informed consent per the guidelines of the Ethics Review Committees of Sri Lanka. Personal identifiable information was not extracted nor disclosed. Approval was obtained from the Head of Military Hospital Narahenpita to conduct the study as an Institutional Review Board has not yet been established.

With the objective of developing a rapid and simple clinical tool for grading traumatic extremity ischemia in combat casualties we formulated the modified Kornilov extremity ischemic index (MKEII). ^9^ (Table I) The MKEII was applied retrospectively to the single surgeon database of 129 extremity vascular injuries that were managed at the Role 3. Patients with isolated iliac artery injury (2), isolated venous injuries (11), deaths due to non-ischemic causes (2) and patients with inadequate data (25) were excluded to form a study cohort of 89 patients. These injuries were assigned to three categories based on the presence of palpable pulse, dopplerable pulse and ankle mobility on presentation to the Role 3. We analyzed the cases in each MKEII category for anatomic distribution, severity of arterial injury, concomitant venous injury, skeletal trauma, damage control adjuncts used and outcomes. Limb salvage was defined as clinically well perfused limbs with documented palpable pulses and/or positive doppler exam at the time of discharge. All amputations performed 24 hours after initial operation were documented as secondary amputations. Vessels with total or segmental loss of more than two-thirds the circumference of the vessel wall were classified as severely injured after analyzing surgeons’ drawings and digital photography taken at the time of surgical exploration.

**Table I.** Modified Kornilov extremity ischemic index (MKEII)

| Category | Pulse | Arterial Doppler | Ankle movements |
| --- | --- | --- | --- |
| C1 | +/- | + | Active |
| C2 | - | - | Passive |
| C3 | - | - | None (fixed) |

The primary objective was to demonstrate the impact of graded ischemia on limb outcome and to validate MKEII based on the Sri Lankan Civil War experience. The secondary objective was to identify specific injury profiles leading to rapid onset of irreversible muscle injury and amputation.

As none of these variables were distributed normally, we used median and interquartile ratio (IQR) for the numerical variables and Mann-Whitney U test and Kruskal Wallis test to compare them. We used Pearson Chi square and Fisher’s exact test to assess the associations between categorical variables. The level of significance was set at alpha 0.05 with a two-tailed test. Statistical analysis was performed using IBM SPSS Statistics. (IBM Corp. Released 2011. IBM SPSS Statistics for Windows, Version 20.0. Armonk, NY: IBM Corp.)

## Results

Based on the MKEII, the 89 arterial injuries were divided into category 1 (C1), category 2 (C2) and category 3 (C3) based on their presenting signs. (Table 2) The majority of the patients were labeled C1 with 54/89 (61%). 24/89 (27%) of the patients presented as C2 and 11/89 (12%) presented as C3. As shown in Table II, there were no amputations in the C1 group, C2 had a 5/24 (21%) amputation rate and C3 had a 7/11 (64%) amputation rate. Chi-square test analysis of these categorical variables revealed a statistically significant P <0.001 correlation.

**Table II.** - Comparison of limb salvage by MKEII classification

| MKEII Classification | Salvaged (n=77) | Amputated (n=12) | P value |
| --- | --- | --- | --- |
| C1 | 54 | 0 | **<0.001\*** |
| C2 | 19 | 5 | **<0.001\*** |
| C3 | 4 | 7 | **<0.001\*** |

\*Chi square Test

Further subgroup analysis compared multiple factors as distributed among the C1, C2 and C3 classification. (Table III) These factors were then independently correlated with amputation versus limb salvage. (Table IV) The distribution of popliteal injuries, severe arterial injuries and concomitant venous injuries were statistically significantly correlated with the C1, C2 and C3 groups. Skeletal injury, amount of blood transfused, time from injury to presentation at Role 3 (representing ischemic time) were not found to correlate with the C1, C2 and C3 groups. When these parameters were compared with the outcome of limb salvage versus amputation, the presence of popliteal injury, concomitant skeletal injury and venous injury, and severe arterial injury were all statistically significantly correlated with amputation. Ischemic time and amount of blood transfused were not correlated with amputation.

**Table III.** – Comparison of clinical parameters per MKEII classification

| Parameter | C1<br>(n=54) | C2<br>(n=24) | C3<br>(n=11) | P value |
| --- | --- | --- | --- | --- |
| Popliteal | 13 | 10 | 8 | **0.006** * |
| Fracture | 11 | 10 | 5 | 0.073* |
| Severe<br>Arterial<br>injury | 28 | 18 | 10 | **0.018** * |
| Concomitant<br>venous injury | 11 | 12 | 8 | **0.001** * |
| Fasciotomy | 25 | 14 | 9 | 0.147* |
| Units blood<br>transfused<br>(Median<br>(IQR)) | 2.0(1.0-3.0) | 2.0 (1.0-4.0) | 3.0(2.5-6.0) | 0.227<sup>†</sup> |
| Transfer<br>time in<br>minutes<br>(Median<br>(IQR)) | 278 (224-371) | 300 (225-378) | 280 (225-630) | 0.905<sup>†</sup> |
| Salvaged | 54 | 19 | 4 | **<0.001** * |

\*Chi square Test
<sup>†</sup>Kruskal Wallis test

**Table IV:** Comparison of clinical parameters in salvaged versus amputated group

| Parameter | Salvaged<br>(n=77) | Amputated<br>(n=12) | P value |
| --- | --- | --- | --- |
| Popliteal | 21 | 10 | **<0.001<sup>\*</sup>** |
| Fracture | 19 | 7 | **0.035<sup>\*</sup>** |
| Severe Arterial injury | 45 | 11 | **0.028<sup>\*</sup>** |
| Concomitant severe<br>venous injury | 20 | 11 | **<0.001** |
| Fasciotomy | 39 | 9 | 0.186<sup>*</sup> |
| Blood units transfused<br>(Median (IQR)) | 2.0 (1.0-4.0) | 3.0 (3.0-3.0) | 0.452<sup>†</sup> |
| Delay in presentation<br>to Role 3 in minutes<br>(Median (IQR)) | 300 (232-385) | 252 (205-270) | 0.164<sup>†</sup> |

\*Chi square Test
<sup>†</sup>Mann Whitney U test

## Discussion

In the combat setting, the critical decision to attempt limb salvage is impacted by the tiered medical care system and is influenced by resources available.^10,11^ Extremities mangled beyond perceptible reconstruction need to be primarily amputated particularly in the setting of mass casualty.^12^ (Figure 1) The remainder of extremity vascular injuries should be correctly identified and triaged for damage control vascular adjuncts. Currently there is no reliable tool to stratify degree of ischemia to decide on the most suitable management plan at the resource limited Role 2 setting. (Figure 2) MESS has been assessed for credibility to be applied to make decisions in the battlefield by multiple authors, however they did not come to a consensus.^7,13-15^

**Figure 2:**
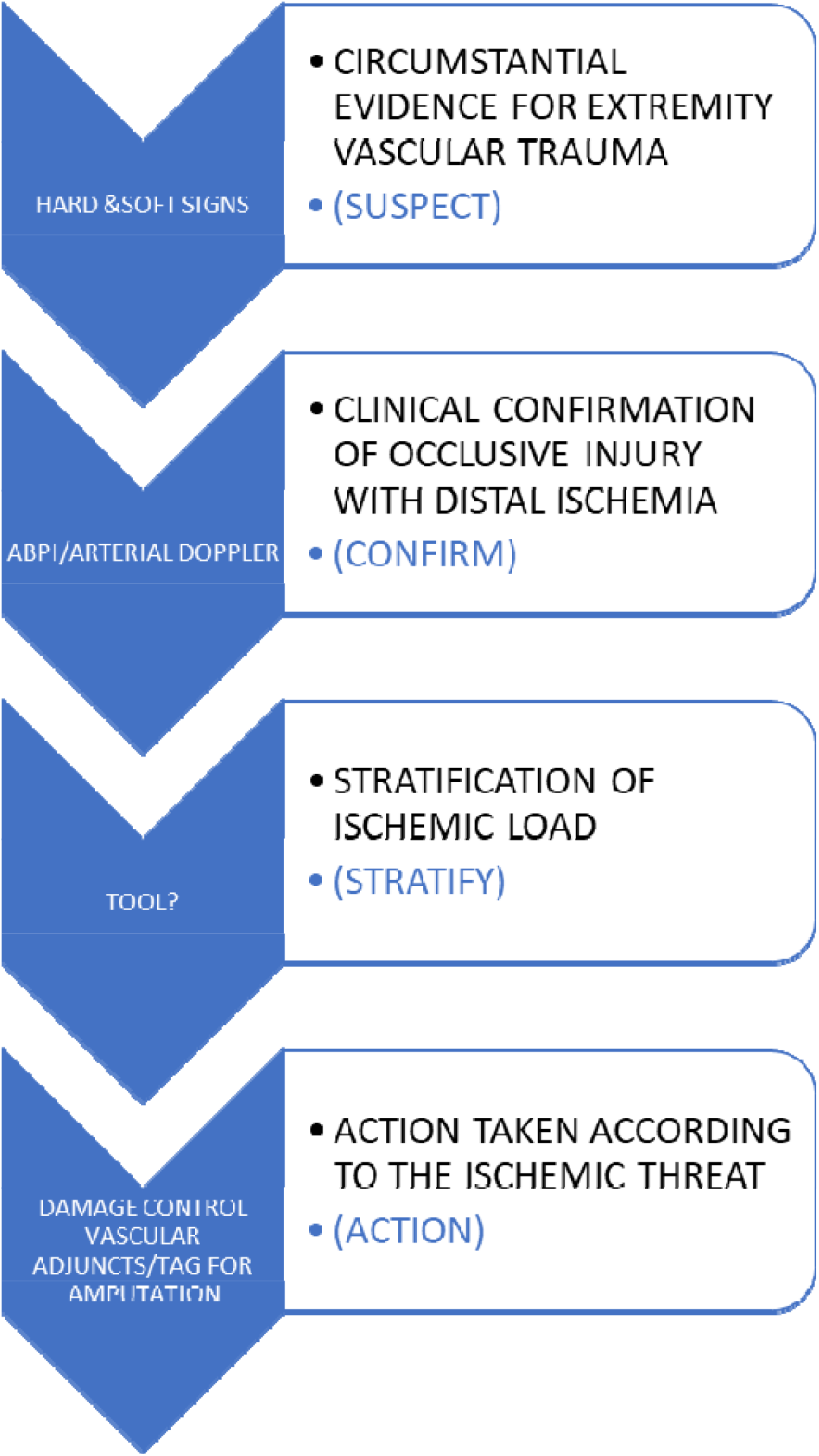
Flow chart for Role 2 combat vascular extremity management.

In extremity vascular injury, there is a spectrum from palpable pulses to ischemic muscles which parallels the degree of main vessel damage and surviving collateral circulation. Accuracy of muscle response to electrical stimulation in the evaluation of muscle viability can be difficult to assess with inhomogenous distribution of ischemia.^16^ The MKEII was creating by adapting the Rutherford classification and the V.A. Kornilov extremity ischemic index to address these issues.^9,17^ In 1971, Kornilov mooted the idea that sensory and motor deficit is partly due to the ischemia in a vascular injured extremity and not due to primary nerve damage. He proposed a simplified grading ischemic index divided into compensated, uncompensated, and irreversible ischemia for peripheral arterial injuries.^18^ The Rutherford classification was created in 1986 as a standard to analyze and report on acute ischemic event in patients with occlusive peripheral arterial disease. The previous categories were further stratified into marginally or early threatened ischemia and immediately threatened or late threatened ischemia bordering with irreversible. We simplified the existing clinical signs to be more practical for the combat setting, namely presence of palpable pulses, audible Doppler signal and ankle rigidity.^19^ Ankle rigidity has been identified by multiple authors as a marker of poor outcome since the Vietnam War era although few have investigated its validity for ischemia grading.^20^ The suggested utility for the MKEII is to triage C1 as delayed, C2 as immediate and C3 as amputation. Although there were 4 extremities salvaged in the C3 group of our study, 3 of these patients had a prolonged hospital course and we have no information about long term function. We suggest that C3 be considered for immediate amputation particularly with mass casualty scenario or limited resources. (Figure 3) C3 patients need further study for resultant functionality of the salvaged limb.

**Figure 3:**
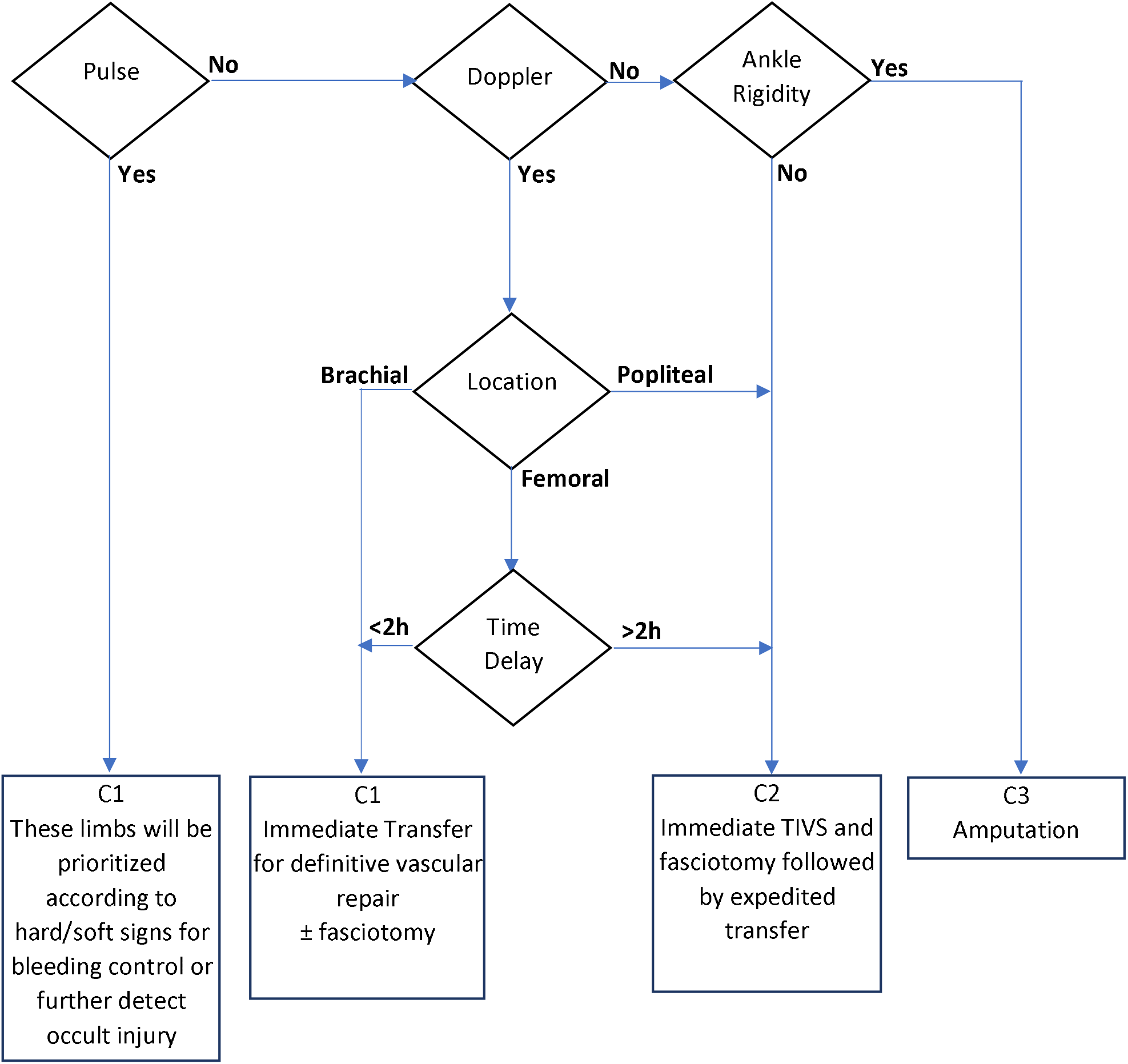
Proposed algorithm for Role 2 combat vascular extremity decision making.

Future research should aim at identifying simple and practical biochemical or imaging modality to stratify the degree of muscle ischemia to near perfection to further facilitate decision making.

Further refining the relationship between ischemic burden and time in extremity vascular injury will aid triage in the battlefield.

This retrospective study is limited by the exclusion of 25 cases excluded for no recorded data, 10 of which underwent immediate amputation. The number of total amputations in our study group is relatively small and this may impact the final data. We lack long-term follow-up to validate ischemia classification for the functionalality aspect of limb salvage.

## Conclusion

By application of the MKEII, combat surgeons can more rapidly and accurately triage extremity vascular injuries to maximize resource utilization to achieve realistic limb salvage goals.

## Data Availability

Available on request

## Disclaimer

The contents of this publication are the sole responsibility of the author(s) and do not necessarily reflect the views, opinions or policies of Uniformed Services University of the Health Sciences (USUHS), the Department of Defense (DoD), the Departments of the Army, Navy, or Air Force. Mention of trade names, commercial products, or organizations does not imply endorsement by the U.S. Government.

## References

1. Kauvar DS, Sarfati MR, Kraiss LW. National trauma databank analysis of mortality and limb loss in isolated lower extremity vascular trauma. J Vasc Surg. 2011;53(6):1598–1603. doi:10.1016/j.jvs.2011.01.056

2. Clouse WD, Rasmussen TE, Peck MA, Eliason JL, Cox MW, Bowser AN, et al. In-theater management of vascular injury: 2 years of the Balad Vascular Registry. J Am Coll Surg. 2007;204(4):625–632. doi:10.1016/j.jamcollsurg.2007.01.040

3. Fortuna G, DuBose JJ, Mendelsberg R, Inaba K, Haider A, Joseph B, et al. Contemporary outcomes of lower extremity vascular repairs extending below the knee: A multicenter retrospective study. J Trauma Acute Care Surg. 2016;81(1):63–70. doi:10.1097/TA.0000000000000996

4. Topal AE, Eren MN, Celik Y. Lower extremity arterial injuries over a six-year period: outcomes, risk factors, and management. Vasc Health Risk Manag. 2010;6:1103-1110. doi:10.2147/VHRM.S15316

5. Liang NL, Alarcon LH, Jeyabalan G, Avgerinos ED, Makaroun MS, Chaer RA. Contemporary outcomes of civilian lower extremity arterial trauma. J Vasc Surg. 2016;64(3):731–736. doi:10.1016/j.jvs.2016.04.052

6. Simmons JD, Gunter JW, Schmieg RE, Manley JD, Rushton FW, Porter JM, et al. Popliteal artery injuries in an urban trauma center with a rural catchment area: do delays in definitive treatment affect amputation? Am Surg. 2011;77(11):1521–1525.

7. Ratnayake A, Bala M, Worlton TJ. The Dilemma of using MESS to Identify Predictors of Poor Outcomes in Extremity Vascular Trauma. J Vasc Surg. Published online April 4, 2020. doi:10.1016/j.jvs.2020.01.081

8. Ratnayake AS, Samarasinghe B, Wijeyaratne M, Sheriffdeen AH. 26 - Asia: Sri Lanka. In: Rasmussen TE, Tai NRM, eds. Rich’s Vascular Trauma (Third Edition). Elsevier; 2016:287–292. doi:10.1016/B978-1-4557-1261-8.00026-6

9. Samokhvalov IM, Pronchenko AA, Reva VA. 29 - Europe: Russia. In: Rasmussen TE, Tai NRM, eds. Rich’s Vascular Trauma (Third Edition). Elsevier; 2016:301-308. doi:10.1016/B978-1-4557-1261-8.00029-1

10. Rasmussen TE, Clouse WD, Jenkins DH, Peck MA, Eliason JL, Smith DL. Echelons of Care and the Management of Wartime Vascular Injury: A Report From the 332nd EMDG/Air Force Theater Hospital, Balad Air Base, Iraq. Perspect Vasc Surg Endovasc Ther. 2006;18(2):91–99. doi:10.1177/1531003506293374

11. Jenkins DH, Tai NRM, Brohi K. 3 - Systems of Care in the Management of Vascular Injury. In: Rasmussen TE, Tai NRM, eds. Rich’s Vascular Trauma (Third Edition). Elsevier; 2016:21-27. doi:10.1016/B978-1-4557-1261-8.00003-5

12. Ratnayake AS, Worlton TJ. Bleeding control in combat fields with extreme transfer time. BMJ Mil Health. 2020;166(3):203–203. doi:10.1136/jramc-2018-001120

13. Brown KV, Ramasamy A, McLeod J, Stapley S, Clasper JC. Predicting the need for early amputation in ballistic mangled extremity injuries. J Trauma. 2009;66(4 Suppl):S93-97; discussion S97-98. doi:10.1097/TA.0b013e31819cdcb0

14. Sheean AJ, Krueger CA, Napierala MA, Stinner DJ, Hsu JR, Skeletal Trauma and Research Consortium (STReC). Evaluation of the mangled extremity severity score in combat-related type III open tibia fracture. J Orthop Trauma. 2014;28(9):523–526. doi:10.1097/BOT.0000000000000054

15. Perkins ZB, Yet B, Sharrock A, Rickard R, Marsh W, Rasmussen TE, et al. Predicting the Outcome of Limb Revascularization in Patients With Lower-extremity Arterial Trauma: Development and External Validation of a Supervised Machine-learning Algorithm to Support Surgical Decisions. Ann Surg. 2020;Publish Ahead of Print. doi:10.1097/SLA.0000000000004132

16. Labbe R, Lindsay T, Walker PM. The extent and distribution of skeletal muscle necrosis after graded periods of complete ischemia. J Vasc Surg. 1987;6(2):152–157. doi: 10.1067/mva.1987.avs0060152

17. Rutherford RB, Baker JD, Ernst C, Johnston KW, Porter JM, Ahn S, et al. Recommended standards for reports dealing with lower extremity ischemia: revised version. J Vasc Surg. 1997;26(3):517–538. doi:10.1016/s0741-5214(97)70045-4

18. Samokhvalov IM, Zavrazhnov AA, Kornilov EA, Margarian SA. [Surgical strategy for combined gun-shot wounds of extremities with injuries of main arteries]. Vestn Khir Im II Grek. 2006;165(5):45–49.

19. Ratnayake AS, Worlton TJ. Pragmatism in Staged Combat Extremity Vascular Management: A Practical Alternative to the Rutherford Classification for Acute Limb Ischemia. Ann Vasc Surg. Published online January 21, 2020. doi:10.1016/j.avsg.2020.01.010

20. Hughes CW. The Primary Repair of Wounds of Major Arteries. Ann Surg. 1955;141(3):297–303.

